# INDI – Integrated Nanobody Database for Immunoinformatics

**DOI:** 10.1101/2021.08.04.21261581

**Authors:** Piotr Deszyński, Jakub Młokosiewicz, Adam Volanakis, Igor Jaszczyszyn, Natalie Castellana, Stefano Bonissone, Rajkumar Ganesan, Konrad Krawczyk

## Abstract

Nanobodies, a subclass of antibodies found in camelids, are a versatile molecular binding scaffold composed of a single polypeptide chain. The small size of nanobodies bestows multiple therapeutic advantages (stability, tumor penetration) with the first therapeutic approval in 2018 cementing the clinical viability of this format. Structured data and sequence information of nanobodies will enable the accelerated clinical development of nanobody-based therapeutics. Though the nanobody sequence and structure data are deposited in the public domain at an accelerating pace, the heterogeneity of sources and lack of standardization hampers reliable harvesting of nanobody information. We address this issue by creating the Integrated Database of Nanobodies for Immunoinformatics (INDI, http://research.naturalantibody.com/nanobodies). INDI collates nanobodies from all the major public outlets of biological sequences: patents, GenBank, next-generation sequencing repositories, structures and scientific publications. We equip INDI with powerful nanobody-specific sequence and text search facilitating access to more than 11 million nanobody sequences. INDI should facilitate development of novel nanobody-specific computational protocols helping to deliver on the therapeutic promise of this drug format.

## 1. Introduction

Antibodies are proteins capable of recognizing a specific molecular site on a potentially noxious molecule (antigen), starting an immune response (1). Because of their binding malleability they are the primary class of biotherapeutics (six of ten blockbusters and market worth ∼100b$). Clinical development of an antibody-based drug is complex and arduous, often taking years (2, 3). The difficulties stem from the complexity of antibodies: they are composed of two polypeptide chains which need to be co-engineered and co-expressed. The protein itself is large which makes delivery difficult especially in challenging cases such as tumor penetration. Therefore there is a lot of interest in exploring alternative antibody formats with more favorable therapeutic properties. One of these is a subclass of antibodies discovered in camelids - the nanobody (alternatively called the single domain antibody or VHH) (4).

Nanobodies bear similarity to normal antibodies however their antigen binding region is composed of just one polypeptide chain. Nanobodies retain molecular recognition advantages of antibodies and exhibit improved biophysical and therapeutic properties as a result of their smaller size (5). Nanobodies are reported to be more stable, soluble and able to recognize cryptic epitopes and penetrate tissues inaccessible to normal antibodies (4, 6). The interest in this direction is reflected by multiple novel nanobodies in either regulatory filing or in the late clinical-trial stages (7) and an increasing volume of patents reporting nanobody sequences (8). In 2018 the first nanobody drug was approved (Caplacizumab (9), by Ablynx), confirming the therapeutic viability of such molecules. Developing nanobodies using traditional laboratory approaches will still require years before they reach the clinic. Computational approaches could accelerate this process, delivering life-saving therapeutics faster and make them more affordable.

Computational methods to design antibodies are already mature enough to provide value in monoclonal antibody therapeutic pipelines (10). By contrast, though nanobodies were discovered close to 30 years ago (11), they attracted less attention in collating data and developing computational protocols addressing these molecules (10). Development of approaches enabling computational design of nanobodies rely on ever deeper analysis of their sequence diversity (12, 13) structural conformations (14), antigen-binding preferences (15), attempts at modifying their binding mode (16) and emerging deep-learning methods tackling this format (17).

Successful computational protocols addressing nanobodies rely on sound sequence and structure data describing the biology of these molecules. A pioneering effort in this direction was achieved by the iCAN (18) and sdAB-DB (19) databases that to our knowledge were first attempts at collection of nanobody-related data. These databases focused on manual identification of antibodies. As a result, they hold a relatively small number of publicly available nanobody data, with sd-AB reporting 1,452 sequences and iCAN 2,391. Data collection frameworks need to keep up pace with the ever-increasing amount of biological sequence data in the public domain. To tackle this, we created INDI-Integrated Nanobody Database for Immunoinformatics. INDI is a novel nanobody database that collates nanobody information from all major data repositories in the public domain, chiefly in automated fashion.

## DATA COLLECTION

We identified five major sources of biological sequence information: NCBI GenBank (20), Protein Data Bank (21), patents (8), next-generation sequencing (NGS) repositories (22, 23) and scientific publications. These sources provide a good coverage associated with systematic repositories collecting protein information from scientific literature and patent documents.

Because of the heterogeneity of the sources, we take the variable sequence of the nanobody as the common denominator between the datasets. We require the nanobody sequences to have all three Complementarity Determining Regions (CDRs) present and only contain 20 canonical amino acids. Sequences are linked with metadata specific for the source dataset (Table 1).

**Table 1.** Contents of INDI in May 2021. Data in INDI is divided into five distinct sources. For each source we provide the reference to the online resource we obtained the data from (with the exception of scientific publications), metadata associated with accessions in source as well as May 2021 statistics of the number of nanobodies we extracted.

| Source | Unique | Unique | Main source | Metadata |
| --- | --- | --- | --- | --- |

|  | **sequences** | **Accessions** |  |  |
| --- | --- | --- | --- | --- |
| Structures | 476 | 702 PDB codes | Protein Data Bank (21) | PDB code, PDB title, authors, resolution technique, text headers of chain fasta files |
| NCBI GenBank | 1768 | 1857 GenBank ids | GenBank (20) | GenBank ID, GenBank description/definition, reported organism, date, reference title reference link/pmid, reference journal, reference authors |
| Next-generation sequencing | 11,228,600 | Seven Bioproject ids | Sequence Read Archive (23) | BioProject id, SRA id |
| Patents | 13,950 | 659 patent families | Patented Antibody Database (8) | Patent number, applicants, patent title, patent abstract |
| Manual | 996 | 91 papers | Scientific publications | Publication title, publication abstract, publication link |

Three of the datasets (PDB, GenBank and patents) are suitable for automatic curation of the data (Figure 1). Here, sequence entries are firstly analyzed for presence of antibodies employing Hidden Markov Models trained on antibody germline genes (24). Sequences where antibodies are detected, are further filtered for presence of nanobodies and thus inclusion in INDI. Accessions containing nanobodies are identified by natural language processing. Through analysis of nanobody-related keywords used in previous studies and our own iterative analysis of nanobody accessions we created a set of keywords relating to nanobodies: *vhh, nanobody, single domain antibody, domain antibody, single variable domain*. Arbitrary pieces of text from our heterogenous sources were normalized by case-folding and stemming and then checked for inclusion of the said keywords.

**Figure 1.**
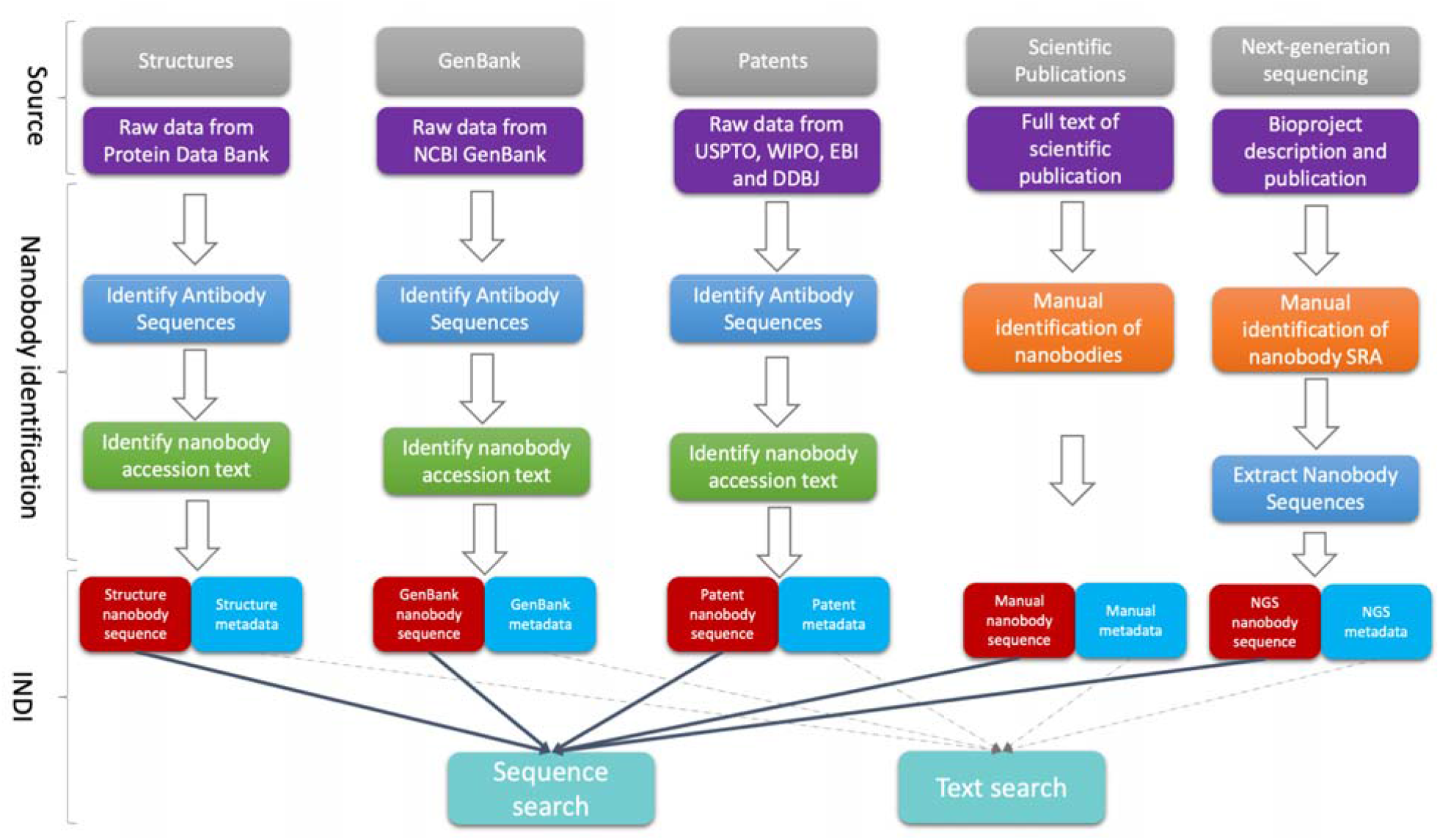
Data sources and information organization in INDI. We obtain nanobody data from five distinct sources: structures, GenBank, patents, scientific publications and NGS. Structures, GenBank and patents are suitable for automated identification divided into identification of antibody sequences and subsequent filtering of nanobody sequences based on text. Scientific publications and NGS are not suitable for automated identification and they require ad-hoc manual curation. Data from all sources are standardized into sequence and metadata indexes. The web-utility of INDI enables users to query nanobody sequence and metadata indexes spanning all five repositories.

NGS and scientific papers are curated manually because of limited standardization. Though NGS depositions are increasingly standardized as a result of the AIRR community efforts (25), specific formats such as nanobodies need to be treated in an ad-hoc manner. For instance, NGS study descriptions need to be manually checked for inclusion of nanobodies so as to avoid errors where single domain antibodies are deposited alongside canonical antibodies (12). Scientific publications containing nanobodies are identified an ad-hoc manner and nanobody sequences found by a human curator included in INDI as part of the ‘manual’ dataset.

In May 2021, INDI held more than 11 million nanobody sequences spanning our five data sources (Table 1). Contents and specific data collection strategies for each of the five datasets making up INDI are described below.

### NCBI GenBank

We collected the protein sequences from the ftp facility of NCBI GenBank in February 2021. Protein sequences were identified as translation entries associated with each GenBank entry. We discarded sequences that were either too short (<70 amino acids long) or too long (>600 amino acids long), with both numbers chosen to capture the lengths of the nanobody variable regions. In total 16,271,610 entries matched these criteria. Antibody sequences were identified employing our adapted version of antibody numbering software (8, 24).

Individual GenBank entries were identified as containing nanobodies if their textual description contained a set of *nanobody-specific keywords* (19, 26) and if the declared organism matched one of: ‘Lama glama’, ‘Camelus dromedarius’, ‘Vicugna pacos’, ‘synthetic construct’, ‘Camelus bactrianus’, ‘Camelidae’ or ‘unidentified unclassified sequences’. Employing this protocol, in May 2021, we identified a total of 1,768 unique variable nanobody sequences from a total of 1857 GenBank accessions.

Each sequence from GenBank is associated with text metadata specific to this repository. These entries were GenBank ID, GenBank description/definition, reported organism, date, reference title, reference link/pmid, reference journal and reference authors.

### Patents

We extracted patent nanobody sequences from our Patented Antibody Database (8) in February 2021. Patent families containing nanobodies were identified by virtue of their classifications C07K2317/569 (Single domain, e.g. dAb, sdAb, VHH, VNAR or nanobody®), C07K2317/22 (from camelids, e.g. camel, llama or dromedary) or if the title and abstract contained one *nanobody-specific keywords*. A Total of 1,013 patent families satisfied the nanobody keyword or category requirements.

In certain cases, claims are laid not only to nanobody sequences within a patent document but also to canonical antibodies. Such families are characterized by a mix of heavy chains aligning to camelid and non-camelid germlines and presence of light chains. Total of 354 families contained both nanobodies and heavy chains of canonical antibodies. Without manual curation it was impossible to tell whether the individual non-camelid heavy chains could function as antibodies on their own. So as to maximize the precision of automated identification, in INDI we only retain sequences where the document declares presence of nanobodies and we only detect heavy chain sequences aligning to camelid germline genes. This resulted in a total of 13,950 unique variable region sequences from 659 patent families.

Each nanobody patent sequence is associated with metadata from the original document. These are patent number, the applicants (e.g. company), patent title and abstract.

### Structures

We sourced nanobody structures from the Protein Data Bank (PDB) ftp facility (21). Antibody sequences from the PDB were identified according to the protocol of the Structural Antibody Database (SAbDab (27)). We excluded sequences that had noncanonical amino acids in their sequences (e.g. 1I3U, 6ULF, 6ANA or 6VLN). From such a constrained set of antibodies, structures containing nanobodies were identified by inclusion of *nanobody-specific keywords* in text associated with the PDB, specifically descriptions of the chains.

Such targeted approach was necessary to distinguish between cases where canonical antibodies might be present alongside nanobodies (e.g. 6ZCZ, 7JOO). Furthermore, it allows us to weed out cases such as 6QKD that reports a VHH-based antibody or 4O9H, which is a camelid Fab rather than a nanobody. Here we also detect human-only single domain antibodies such as 5N88. In May 2021 this extraction approach identified 476 unique nanobody chains from 702 PDBs.

Each structural nanobody entry is linked to metadata originating from the PDB accession. These entries are the PDB code, accession title, authors, resolution, technique (e.g. x-ray) and text headers of FASTA files associated with individual chains.

### Next Generation Sequencing

We identified eight bioprojects reporting next-generation sequencing of nanobodies by employing the text-search utility at NCBI: PRJDB2382, PRJEB7678, PRJNA642677, PRJDB7792, PRJEB25673, PRJNA516512, PRJNA638614 and PRJNA321369. We extracted the nanobody sequences contained within the bioprojects as described previously (22). In brief, SRA files containing raw reads are analyzed using IgBlast (28) that translates the nucleotide sequences into amino acids. Sequences that are free of stop codons and that contain all three CDRs are retained.

In the case of the Bactrian camel study that contained samples of both nanobodies and canonical antibodies, we only made the VHH samples part of INDI that corresponded to SRAs SRR3544218, SRR3544220 and SRR3544222. Though, PRJNA516512 advertised presence of nanobodies, the resulting IgBlast processed sequences did not satisfy our inclusion criteria due to incompleteness of chains. The remaining seven bioprojects contributed a total of 11,228,600 unique variable region sequences. Each NGS nanobody sequence is associated with the SRA file it originated from and metadata of the Bioproject.

### Manual curation

In certain cases individual nanobody sequences that are reported in publications are not deposited in systematic repositories such as GenBank or the PDB. Such sequences are typically contained in supplementary materials. The reporting is not standardized and therefore challenging for automated approaches. Therefore, we created a ‘manual’ category for all sequences that are obtained by human curation of sequences originating directly from scientific publications. Because of lack of automated means, data in this database will be updated in an ad-hoc way. In each case, the variable region sequences of nanobodies are manually identified by a human curator and automatically filtered for presence of all three CDRs and lack of non-canonical amino acids. The variable region sequences are associated with the metadata of the original publication in the form of its title and abstract.

### Comparison to other databases

In order to ensure that we capture nanobody information reliably, we performed a contrast to other resources that collect these molecules. We compared entries from INDI to those in sdAB-DB (20), structural nanobody entries curated by the Structural Antibody Database (SAbDab) (27), and the CoV-AbDab that curates some COVID-19-related nanobodies alongside canonical antibodies (29).

We checked whether all the PDB codes that we identified as nanobodies have the same annotation in SAbDab. All of the February 10th version of SAbDab nanobody-annotated sequences were found in INDI. The only exceptions were PDB codes that were subsequently changed in the PDB (e.g. 6h7k-> 6ibl, 6csy->6mxt). Entries from CoV-AbDab (Mar 17th version) and sdAB-DB that were manually extracted from the literature were not found in INDI. All the entries that could be obtained by automatic means from the PDB or GenBank were shared between INDI, CoV-AbDab and sdAB-DB.

Entries from sdAB-DB that could have been obtained by automatic means but were not present in our database could be divided between light chain single-domain antibodies (e.g. sdAB-DB sdAb_2370_Sy, GenBank AAG49009, AAG49011) or single chain Fv (scFv) fragments that were deposited as separate chains (e.g. sdAb_1934_Sy, GenBank AKR15657). Certain entries that were present in sdAB-DB contained non-canonical amino acids which we do not include in INDI (e.g. sdAb_1339_Lg CAH60929).

All the literature entries that were not found in INDI were subsequently entered.

## USAGE

We mapped the most common retrieval tasks to facilitate interaction with INDI online and offline. Through our website (http://research.naturalantibody.com) users are able to perform nanobody-specific sequence-based searches and metadata retrieval. To facilitate offline immunoinformatic analyses, we make the data available for bulk download.

### Sequence-based search

We make two nanobody-specific sequence search functions available to facilitate interaction with the data in INDI - *Variable Region Search* and *CDRH3* search. The division reflects the two-common use-cases of nanobody sequence identification. The former addresses retrieval of the entirety of the variable region.

The latter addresses specific searches of the most variable portion of the nanobody responsible for most of the antigen-contacts, namely CDRH3.

Variable Region Search addresses retrieval of the entire nanobody sequences that are best matched to the query. In order to reflect the nanobody-specific nature of the search, we compare nanobody sequences using the IMGT scheme, which provides an immunoglobulin-specific framework for alignment of antibodies/nanobodies. The query sequence is IMGT-numbered and subsequently aligned to the pre-numbered nanobody sequences in INDI based on IMGT-positions. The results are sorted by the highest sequence identity over the entire variable region. The results are given in an interactive sortable table that leads to more detailed results on each hit. Users can sort the results by the entire variable region sequence identity as well as the IMGT-identity to individual CDRs.

Of the three-heavy chain CDRs, CDRH3 carries the largest number of antigenic contacts (15) and is often used as a proxy for antigenic specificity by itself. Therefore, we equipped INDI with a search facility retrieving CDRH3, disregarding the rest of the variable region. Input to CDRH3 search is the IMGT-defined sequence of the CDRH3. The sequence is then divided into k-mers with k=4. The query k-mers are matched against these precomputed for each sequence in INDI. The hits are sorted by the number of k-mers in common with the query. Results are subsequently aligned using global pairwise alignment algorithm as implemented in Biopython. This allows for length-independent retrieval of sequence similar CDRH3s matches. The CDRH3 results are presented in an interactive sortable table that allows the user to browse through the results and follow links to variable sequences and their associated metadata.

### Text search

Nanobody sequences in INDI are associated with rich textual annotations revealing among others biological targets, origins and purpose of the study of the molecules. Metadata fields are heterogenous across the sources and within them. For instance, metadata associated with structures will contain specific crystallographic parameters not present in other databases. In GenBank, information about the target of a nanobody can be contained within the description of the accession or the individual translations as there is no standardized way to report such information. The great diversity in text representations poses a challenge in document retrieval.

To address the problem of information retrieval across the five diverse sources, we implemented a text index created on all the metadata fields in all the databases. User is asked to provide the keywords of interest and INDI will retrieve the accessions best matching the results. Users can specify possible targets of nanobodies that are reported as part of the depositions (e.g. protein name VEGF) as well as individual accession numbers (e.g. PDB accession 7JOO).

Results are displayed as an interactive table listing the accessions, source databases and text fields. Users can sort through the results and display the details of matching text entries. The details of text entries are displayed together with nanobody sequences linked to the accession.

### Bulk Download

To supplement our web-based retrieval we make the data available for offline use as well. The data are available as an extract of the two pillars of our data model – sequences and metadata separately. The sequence-extract contains the V-region sequences of nanobodies we identify. Each sequence entry is linked to the metadata fields contained within the meta-extract. Metadata fields are sorted by one of the five databases. All data is available through the main INDI website available at http://research.naturalantibodo.com/nanobodies

## Discussion

Delivering an antibody drug to clinical use requires a big investment of time and resources with a high likelihood of failure at the clinical trials stage. Novel formats such as nanobodies with favorable biophysical properties, offer opportunities to mitigate certain drug discovery risks (5, 30). Innovative approaches for targeted delivery of nanobody-based therapeutics are being pursued currently (31).

Besides the molecular therapies (31), nanobodies are being used in the development of several cellular therapies (31–35). Developing nanobody therapies using traditional lab-based approaches still carries an overhead of many years of experimentation before they reach the clinic. Computational approaches could accelerate this process, delivering life-saving therapeutics much faster.

Though still in its infancy, bioinformatic methods addressing issues of therapeutic nanobody design are being developed. Computational nanobody approaches can provide insight in developing reliable structural modeling methods (36), design of phage display libraries (37) or computational design of novel nanobodies (16). Parallels between antibodies and nanobodies allow certain protocols to transfer information between the two types of molecules. For instance, though nanobody-specific structural modeling pipelines exist (36), it is possible to obtain reliable models of nanobody structures employing antibody protocols (38).

Despite certain parallels between nanobodies and antibodies, contrast between the binding sites of the two reveals certain differences (15). Though there is an overlap between nanobody and antibody epitopes, either is capable of binding molecular surfaces that the other might find challenging (39).

Deconvoluting such nuanced distinctions is required to understand the binding mode of nanobodies ultimately leading to reliable computational nanobody design protocols (16). Any computational efforts however require sound access to nanobody sequence data.

To address this need, here we developed INDI, a database integrating nanobody sequences, structures and their associated metadata in the public domain. Automatic updates from the heterogeneous sources make it possible to keep up with the accelerating pace of deposition in the public domain. Heterogeneity of data in INDI allows nanobody researchers to obtain an accurate picture of the current state of knowledge of nanobody sequence, structure and function. Such knowledge can then accelerate the development of analytical frameworks (14, 15), structural modeling (36), de novo nanobody design protocols (16) and as a basis for deep-learning models addressing nanobody design (17). Altogether we hope that INDI will form a solid data foundation to develop nanobody-specific computational methods that will accelerate development of novel therapeutics in this format.

## Data Availability

All data are available at http://research.naturalantibody.com/nanobodies

